# Addressing a climate emergency amidst the COVID-19 pandemic: A mixed-methods study on a hospital evacuation during the 2021 European floods

**DOI:** 10.1101/2023.02.02.23285386

**Authors:** Willemijn vd Wal, Dennis G. Barten, Linsay Ketelings, Frits v Osch, Madhura Rao, Luc Mortelmans, Joost Bierens

## Abstract

**Introduction:** In July 2021, several European countries were affected by severe floods with water levels of the river Meuse reaching a record high. VieCuri Medical Center (Venlo, the Netherlands) is a hospital located directly adjacent to this river, and in response to the flood threat it was decided to completely evacuate the hospital. The aim of this study was to explore the decision-making process of this emergent evacuation.

**Methods:** A mixed-method approach was used. Qualitative data were collected through semi-structured interviews with 11 key participants closely involved in the evacuation. Quantitative data on the patients that were admitted at the time of the evacuation decision were collected, and included 30-day mortality, 7-day readmission rates and Charlson Comorbidity Index.

**Results:** Three themes were constructed from the interviews: risk-assessment, COVID-19 experience and collaboration. Participants highlighted the role of previous experiences from the COVID-19 pandemic. The use of a national patient coordination center enabled to rapidly assess capacity of potential destination hospitals. Furthermore, the hospital’s preparedness for evacuation could be improved by a thorough analysis of locoregional hazards and preparing for loss of regional healthcare capacity. Findings unrelated to decision-making included the inefficiency of large-scale ambulance dispatches and the expansion of business continuity plans. No patients died unanticipated during this hospital evacuation or within 30 days.

**Conclusion:** Experiences of the COVID-19 pandemic and the availability of a national patient coordination center were found to be decisive in performing this evacuation. This allowed for the swift identification of available capacity in appropriate destination hospitals.

## 1 INTRODUCTION

Hospitals are expected to respond to natural disasters and mass-casualty events. These sudden onset events may severely disrupt the everyday, routine services of a hospital facility and subsequently create a threat to patient care. If this is the case, such events are referred to as ‘internal hospital crises and disasters’ (IHCDs). IHCDs commonly occur and are frequently associated with the evacuation of patients [1]. Previous studies have shown that hospitals’ crisis plans could be improved with regards to evacuation planning [2,3,4,5]. Evacuation plans are often lacking [3,4] not updated, or hardly practiced [4]. Furthermore, hospital crisis plans frequently fail to integrate important aspects such as disaster characteristics, patient numbers and priorities, patient mobility, available staff, available transport and care capacity [2].

Over the past five decades, the worldwide incidence of natural disasters has grown fivefold [6], which is largely caused by an increase of climate-related emergencies [6,7]. Also, the scale of disasters has expanded as a result of increased rates of urbanization, environmental degradation, and intensifying climate variables [7,8]. Hospital locations will not be immune to these global trends. It is therefore likely that hospital evacuations will occur more frequently in the near future [6,7]. In some countries, over 50% of the healthcare facilities are situated in high-risk areas for natural disasters [4]. In numerous high-income countries, including the Netherlands, the majority of the hospitals are located in flood-prone areas [4,9].

In July 2021, several European countries were affected by severe floods [10]. The floods affected several river basins across Europe, including Austria, Belgium, Croatia, Germany, Italy, Luxembourg, the Netherlands and Switzerland [10]. In Belgium and Germany, the floods were catastrophic, causing 221 deaths and widespread damage [10,11]. In the Netherlands, the river Meuse reached its highest summertime level in over 100 years [12]. VieCuri Medical Center (VCMC) is located directly adjacent to this river, in the city of Venlo, which has a population of 85,000 inhabitants. The rising levels of the Meuse resulted in the decision to completely evacuate the hospital on July 16th, 2021.

Hospital evacuations are considered risky operations [9]. Not only because of the transfer of the most vulnerable individuals, but the closure of a hospital (unit) may also result in decreased health care capacity in a wide region [9]. Consequently, the decision to evacuate a hospital is not taken lightly: the anticipated impact on the hospital has to outweigh the risks of the evacuation operation itself.

Limited research has been performed on how critical evacuation decisions are made. One study conducted in Iran identified some important factors, including risk-assessment and estimation, continuing service provision as necessary prerequisites for evacuation [13]. The latter included capacity of hospitals where evacuees can be transferred to, the pathways for evacuation and the availability of adequate ambulances, equipment, staff and communication systems [13]. A study conducted in the mid-Atlantic region on the evacuation decision in response to hurricane Sandy identified the risk to patients’ health as the most decisive factor regarding hospital evacuation, as well as prior experience, costs and continuity of operations [14]. Because data collection on mortality and morbidity associated with previous hospital evacuations would enhance risk-assessments that inform decision-making [14], it would be valuable to also address patient outcomes in relation to the evacuation decision.

This study explores the decision-making process of the hospital evacuation on July 16th, 2021. Mortality and readmission rates were examined to globally assess evacuation-associated patient risks.

## 1. MATERIALS AND METHODS

### 2.1 Setting

VCMC is a teaching hospital located in the province of Limburg in the southeast of the Netherlands. It has two locations: one acute care hospital in Venlo (bed capacity of 396 patients) and one day-care hospital in Venray (bed capacity of 26 patients). The distance between the facilities is 27 km. The adherence area covers a wide area and facilitates 280,000 inhabitants. The hospital in Venlo is located adjacent to the river Meuse. The distance between the Meuse and the hospital in Venray is 8 km. This day-care hospital does not operate an ED and ICU. A satellite photo of VCMC Venlo is shown in Figure 1.

**Figure 1.**
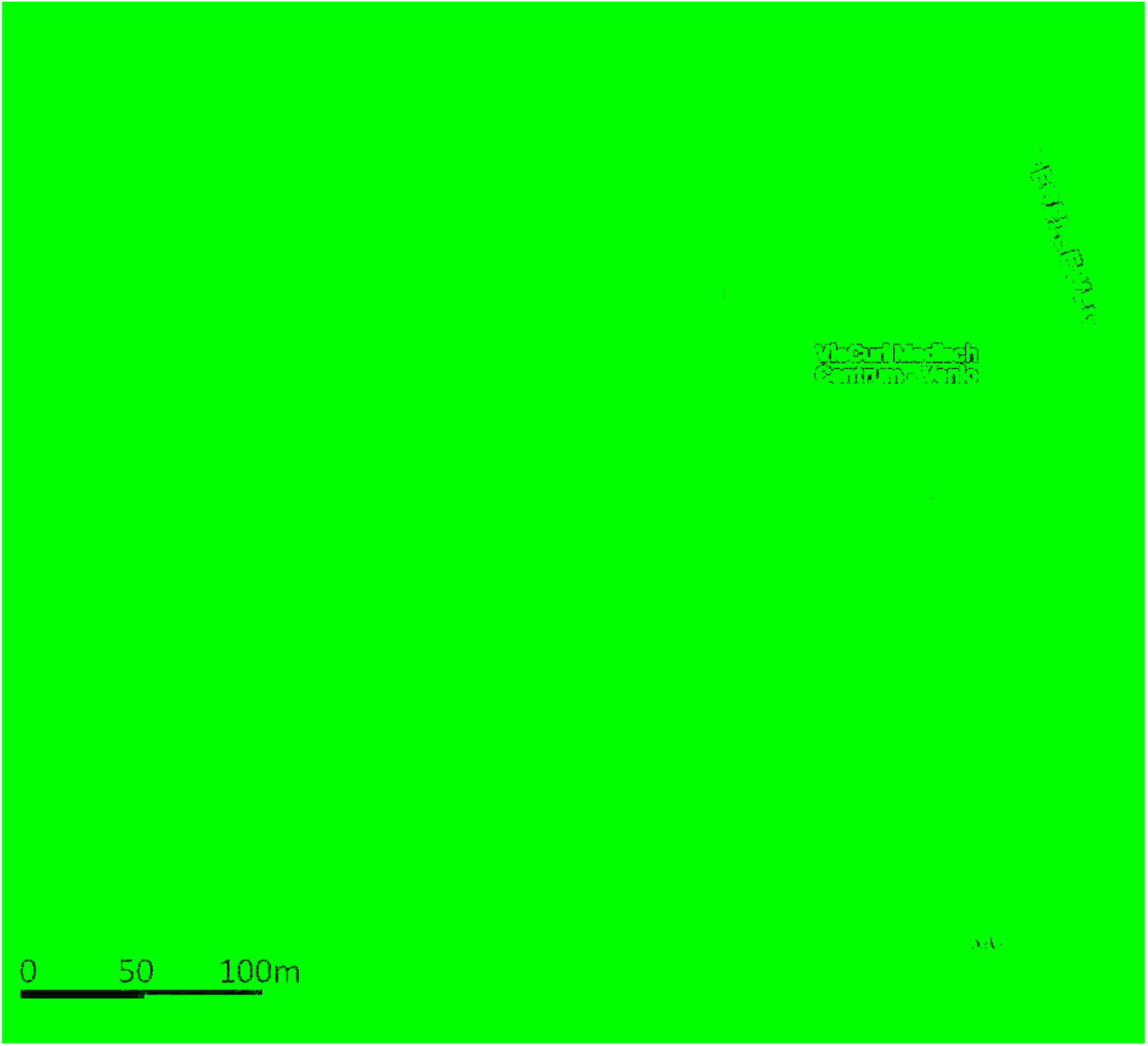
Map of geographical setting of VieCuri Medical Center, Venlo, the Netherlands.

Following heavy rainfall in the catchment area of the river Meuse during several days in July 2021, the water level had been increasing, and on July 15th the local authorities informed the hospital that a further increase was expected. On that same day, the head of the board of directors, who chaired the hospital crisis management team, and the regional safety authority (‘Veiligheidsregio’) decided to completely evacuate the hospital on July 17th. The evacuation decision was communicated one day in advance, on July 16th 10.30 AM. Two hours after this communique, the water level forecasts suddenly changed. The water peak would be reached fifteen hours earlier than expected and the hospital evacuation had to be completed on that same day. The first patient was transferred at 6 PM and the hospital was completely cleared by 11 PM. On July 21st, the hospital was fully operational again and patients were re-transferred to VCMC. The timeline of the evacuation is shown in Figure 2.

**Figure 2.**
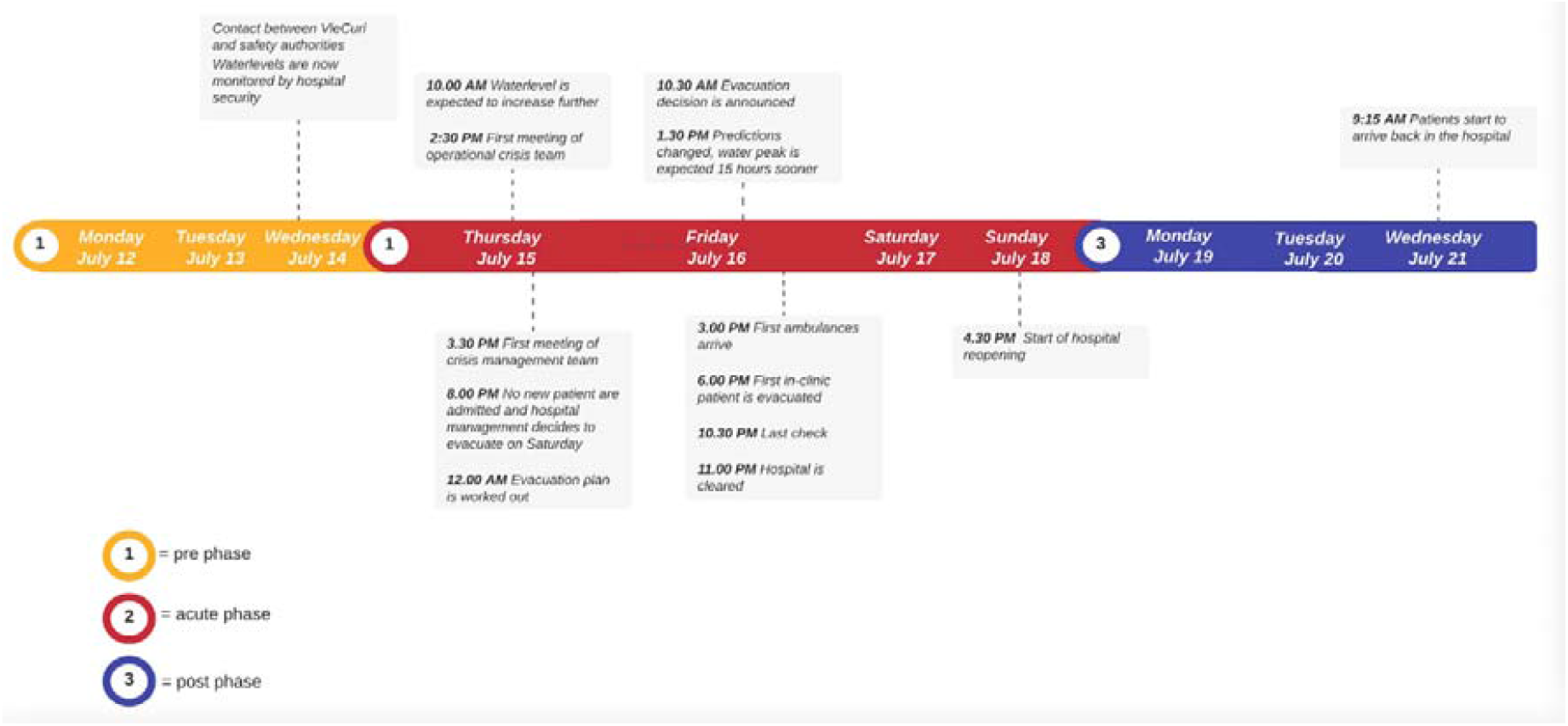
Timeline of the evacuation of VCMC.

The water peak reached the hospital area in the early hours of July 17th but did not surpass the dikes situated along the riverbanks (Figure 3). The hospital remained unflooded and undamaged.

**Figure 3.**
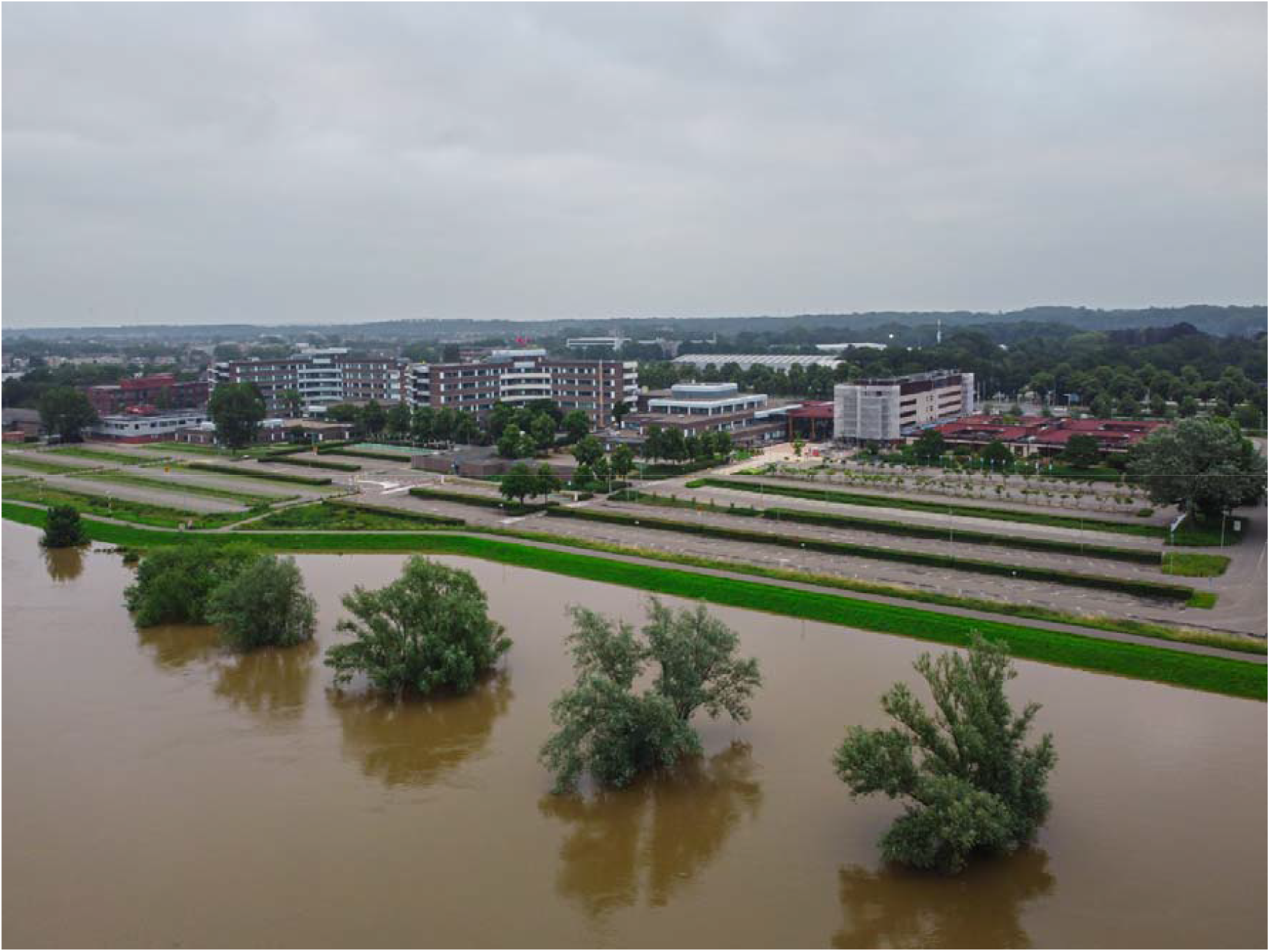
The water level of the Meuse did not surpass the the dikes situated along the riverbanks (Photo by Frank Leenen).

The National Patient Coordination Center (NPCC; Landelijk Coördinatiecentrum Patiëntenspreiding: LCPS) is a Dutch organization established in March 2020 in response to the COVID-19 pandemic. NPCC facilitates the transfer of patients between the different regions of the Netherlands. By distributing patients inter-regionally instead of regionally, it was aimed to evenly divide COVID-19 related workload among hospitals and enhance the continuity of care provision on a national level [15,16].

### 1.2 Study design

A mixed-methods study using a convergent parallel-databases design was performed. In this approach, qualitative and quantitative data were collected and analyzed independently. These were only brought together during the interpretation of the analyzed data [17].

Semi-structured interviews, focused on factors that may have influenced the decision-making process related to the evacuation, were used as qualitative data and explored through thematic content analysis [18]. A descriptive assessment of patient-related outcomes of the patients was used for the quantitative part of this study. This mixed method design allowed for enriching the qualitative findings on decision-making with the outcome of medical indicators of evacuated patients [17]. Methods for the quantitative and qualitative components are described separately. Qualitative data were reported according to the Consolidated Criteria for Reporting Qualitative Research (COREQ) guidelines [19].

### 2.3 Qualitative component

The qualitative component consisted of eleven semi-structured in-depth interviews about the decision-making process, conducted by WvdW [20]. Interview guidelines were constructed, addressing themes based on literature. Initial drafts of the questionnaires were evaluated for validity by the first author (WvdW), one emergency physician (DB), two researchers with experience in qualitative research (LK, MR) and a clinical epidemiologist (FvO). Subsequent drafts were tested in a small sample of the target population to evaluate understandability and feasibility. Based on the test’s findings, the questionnaire was adapted. The final questionnaire included ten open-ended questions. Questions addressed knowledge, feelings and experiences. This allowed for inquiring about participants’ factual information, eliciting emotions and gaining insights on participants’ behavior [21]. The interview guideline can be found in Appendix I. Purposive sampling [20] was used to determine the study population, which included officials in the (crisis) management team of VCMC as well as officials of the regional safety authority working closely with the hospital’s (crisis) managers. NPCC officials were contacted as they had been closely involved in the evacuation process. Semi-structured interviews enabled participants to introduce topics that are of importance to them and allowed for exploring personal insights [20]. The participants were interviewed individually and had no information from the other interviews.

#### 2.3.1 Data collection

Participants were informed and invited per e-mail between October and December 2021, three to five months after the hospital evacuation. The interviews took place at VCMC, the natural working environment of the participants, in the Dutch language. If the participant preferred so, the interview took place digitally using Zoom version 5.8.4. (2421) (Zoom Video Communications, San Jose, CA, USA). Before the interviews commenced, informed consent was signed by the participants, agreeing to the recording and storage of the anonymized material. The interviews were recorded using a voice recorder (DVT1150 Phillips, Amsterdam, the Netherlands).

#### 2.3.2 Data analysis

The interview recordings were transcribed *verbatim* in Microsoft Word version 16.50 (Microsoft Corporation, Redmond, USA) and the transcripts were imported to the coding software Atlas.Ti Mac version 9.1.2 (2087) (Atlas.Ti. Scientific Software Developer GmbH, Berlin, Germany). The process of comparative thematic analysis by means of coding was started after conducting the fourth interview. With open coding, labels were attributed to parts of the texts to give meaning to it [21]. A total number of 55 codes were grouped into 30 categories with axial coding as described by Strauss and Corbin [22], to refine (sub)categories based on connecting codes. Selective coding [22] was used to create overarching categories of the remaining codes. Analysis was ongoing while more interviews were conducted. Quotations that presented the subcategories were then selected and translated from Dutch to English by the first author while ensuring that the original meaning and context were retained. Quotations were returned to participants to check for accuracy. Data saturation was reached when no new themes were drawn from the interviews [23].

### 2.4 Quantitative component

#### 2.4.1 Research design

The quantitative part of the research consisted of a retrospective medical record review of the evacuated patients. The medical record study included all patients admitted to one of the hospital wards on July 16th, 2021 at 10.30 a.m., just before the evacuation decision was internally communicated.

#### 2.4.2 Data collection

Data were collected manually from the electronic medical records by WdW and DB and consisted of demographics (age, sex), date of admission, hospital ward admitted to, primary diagnosis, comorbidities defined by Charlson Comorbidity Index (CCI) [24] and code status (no resuscitation (DNR), no intubation (DNI), or no ICU admission (no ICU). If the patient was evacuated to another healthcare facility, the transfer distance in kilometers was calculated. It was also documented whether patients were discharged prematurely or if their discharge was planned on July 16th 2021. Furthermore, readmission within 7 days and mortality within 30 days after the evacuation event were registered.

#### 2.4.3 Data analysis

Baseline characteristics were analyzed using descriptive statistics on the extracted data, using IBM SPPS Statistics version 25.0 for Windows (IBM Corporation, Armonk N.Y., USA). For age and CCI, medians and interquartile range (IQR) were used. All other data were described in absolute numbers and percentages of patients. The data were used to construct a flow-chart illustrating the mortality and readmission rates amongst the patients, who were grouped according to their placement (transferred to other facility/discharged prematurely/discharged as planned). Demographics and CCI were used to provide insights into the group characteristics.

### 2.5 Ethical considerations

A waiver for ethical approval was provided by the medical-ethical review board of Maastricht University Medical Center (2021-2895) (Appendix III).

## 2. RESULTS

### 3.1 Qualitative results

The sample size was predetermined and consisted of eleven participants. The sample consisted of participants with different functions: hospital crisis-coordinators (n=2), hospital management-team members (of whom also a medical specialist) (n=2), hospital operational-team members (n=2), a medical coordinator of ICU (n=1), medical officials of the regional safety authority (n=2) and medical professionals involved in NPCC (of whom one employee of the hospital that was evacuated) (n=2).The response rate was 100%. Five physical one-on-one interviews took place, six interviews took place digitally. The duration of the interviews varied from 28 to 68 minutes (median: 44 minutes) and data saturation [23] was reached after nine interviews. The final two interviews confirmed that there were no new themes drawn from the data collected.

### 3.2 Themes

Three themes were constructed from the thematic analysis of the semi-structured interviews that focused on the decision-making process: *risk-assessment, COVID-19 experience and collaboration*. The main themes consisted of multiple subthemes (Table 1). In addition, several topics that were not related to the research question were introduced by the participants. These topics are specified under the heading ‘non-decision making related topics’.

**Table 1.**
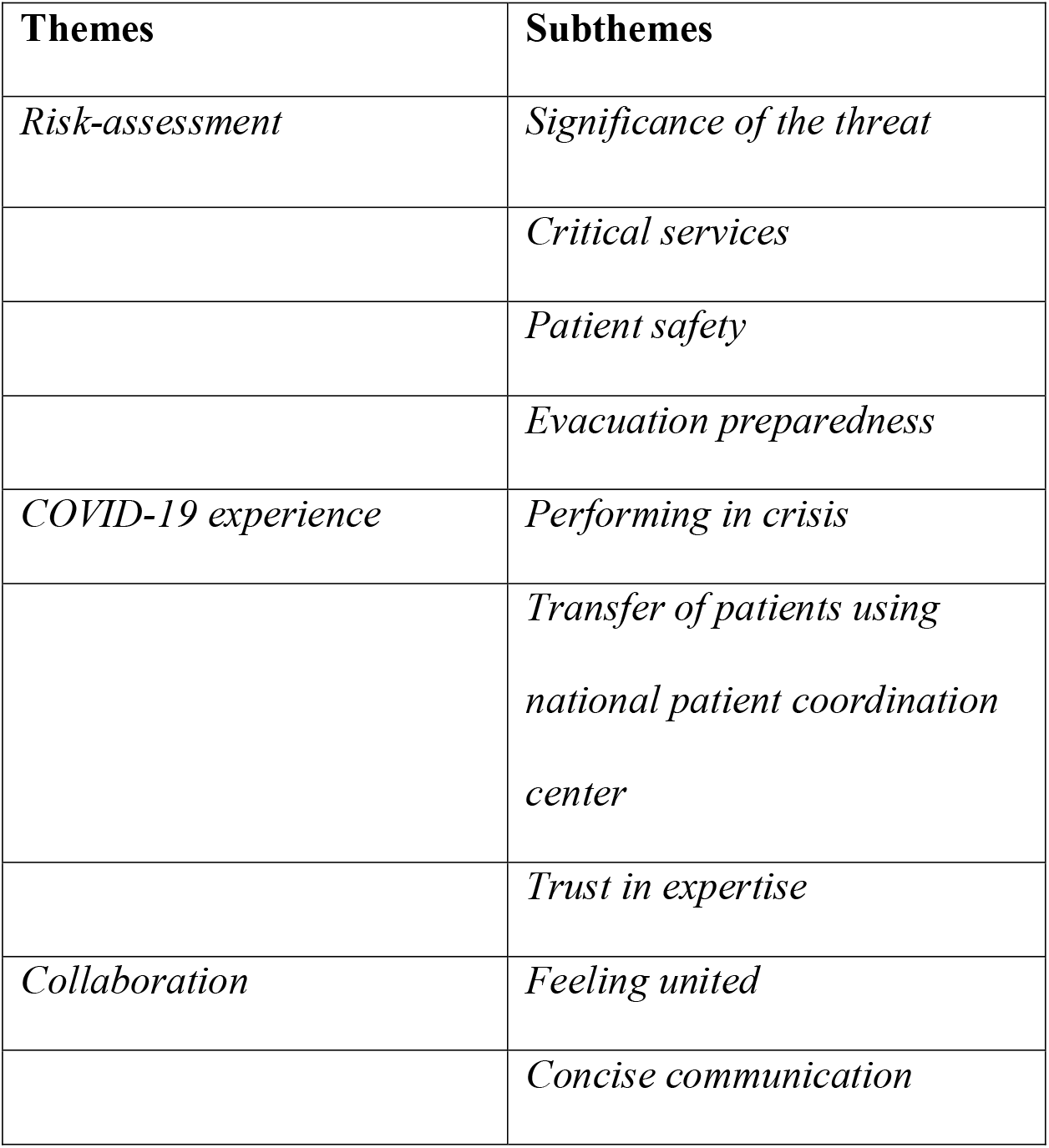
Major themes constructed from semi-structured interviews.

| Themes | Subthemes |
| --- | --- |
| *Risk-assessment* | *Significance of the threat* |
|  | *Critical services* |
|  | *Patient safety* |
|  | *Evacuation preparedness* |
| *COVID-19 experience* | *Performing in crisis* |
|  | *Transfer of patients using national patient coordination center* |
|  | *Trust in expertise* |
| *Collaboration* | *Feeling united* |
|  | *Concise communication* |

#### 3.2.1 Risk-assessment

Participants described the risks they considered and how these risks influenced decision-making. Risks to patient safety and not being able to guarantee healthcare provision led participants to believe that evacuation was the best option. This ultimately resulted in the decision to completely evacuate the hospital.

##### Significance of the threat

Although it was evident that the water level of the river Meuse was rising very rapidly, some participants perceived a lack of information on the significance of the threat. “*You could notice that within the regional safety authority, as well as for example ‘Waterschap’ and ‘Rijkswaterstaat’* [governmental authorities for execution of public works and water management], *those obviously were the most important sources of information, they were unable to point out the effect on the different dike ring areas and where the weak points were located*.*” (Participant 7)*

Participants believed the information was available but not successfully and comprehensively shared with stakeholders. This led to a situation in which the hospital’s crisis management team had to make a decision about evacuation without knowing when the water would reach the area that would affect the hospital and whether the dikes would hold. Most participants reported that this uncertainty had no impact on the evacuation decision. However, it did lead to a lack of understanding of the urgency in which action was required and the timeframe in which the evacuation had to be completed.

##### Critical services

When discussing the anticipated consequences of flooding, participants expressed their concerns about critical services such as power generators, services that are located in the basement and on the ground floor, making them particularly susceptible to failure even when exposed to small amounts of water. *“*…*the hazard was that, even if there would only be 10-12 cm of water, the critical services would stop functioning*.*” (Participant 2)*

Participants stated that it was difficult to establish accuracy on which supplies would be harmed and when. However, if only one of the critical services stops functioning, the safety and continuity of healthcare is at stake.

##### Patient safety

Participants stated they were aware of the risks imposed on patients, especially since the evacuation was a precautionary measure rather than motivated by an immediate threat situation. One participant explained: *“*…*that a patient was getting poor medical care whilst there ultimately was no flood, so you’re taking a precaution, and because you’re taking a precaution you’re putting a patient at risk. We were very aware of that, being the absolute hazard*.*” (Participant 1)*

Well aware of the risks, the majority of the participants never considered the evacuation operation unsafe. The healthcare requirements of every individual patient were carefully considered when planning patient transfers and relocation. Participants believed that adequate healthcare could be provided during transport because of the high quality of the national ambulance services.

##### Evacuation preparedness

Some participants mentioned that the hospital crisis plan specified *when* to evacuate the hospital, but not *how* to do it. Participants noted that every disaster requires a different approach and one can only prepare generic. Therefore, participants thought it might be useful to focus on preventative measures and make risk inventories, e.g. knowing which services are critical, and secure them if possible. Additionally, knowledge should be gained on risks associated with the hospital’s location along the river Meuse.

#### 3.2.2 COVID-19 experience

Participants repeatedly reported that the recent experience with the COVID-19 pandemic played a major role during the flood threat and evacuation. The decision-making process was advanced by this experience too.

> *“The coronavirus crisis, … as the saying goes ‘never waste a good crisis’, you can learn bloody much from it! And yes… you wouldn’t wish any hospital a crisis but requiring all hands on deck sometimes isn’t bad. I think it has its advantages*.*” (Participant 4)*

##### Performing in crisis

Participants experienced benefits from being familiar with their role and the role of others in the crisis management teams that were constructed during the pandemic. Because many functions were fulfilled by the same individuals as during the pandemic, participants experienced almost no uncertainties regarding their function and corresponding tasks. One participant stated that he had learned from performance evaluations after the first COVID-19 wave and now felt more confident to address any uncertainties and to keep colleagues in perspective. Another benefit mentioned was knowing how to take care of patients and how to resume patient care when the crisis came to an end.

##### Transfer of patients using NPCC

Besides their experiences in performing in crises, participants referred to the key role of the NPCC in identifying capacity in appropriate destination hospitals. Although VCMC had already started planning transfers of patients to the nearest hospitals, the NPCC advised otherwise. *“*…*all the ICU patients were already assigned a hospital bed and families were informed that they would be transferred to the nearest hospital. Everyone is happy, physician is happy, recipient is happy, ambulance crew is happy. But then we said, we are not going to do that*.*” (Participant 1)*

Instead of transferring all patients to the nearest hospitals, some patients were purposefully transferred to hospitals over 100 km away. Natural disasters potentially affect multiple healthcare facilities simultaneously and may also lead to a higher healthcare demand in the affected region. Distributing patients inter-regionally is crucial to preserve local emergency care capacity.

The majority of the participants considered the use of NPCC a crucial element in the swift execution of this evacuation operation, and suggest considering a permanent, on-call system, that is here to stay even when the pandemic has ended.

Although confident with the course of events, one participant pointed out there are more ways to get to the same result: *“Maybe the co-occurrence of the pandemic and the foundation of NPCC were just random coincidences, then you should not go around saying we have always done it wrong in the past, because look how it went down in Venlo. It could have been a coincidence but that’s also crisis management, right?” (Participant 10)*

##### Trust in expertise

Many participants felt that officials knew their role and only interfered with matters they had expertise in. Largely, this familiarity was due to the recent experience of the COVID-pandemic.

> *“Simply trust the organization…Everybody’s got their own expertise, if you’ve got faith in that, it will turn out fine*.*” (Participant 8)*

#### 3.2.3 Collaboration

The majority of the participants expressed they had especially valued working together prior to and during the evacuation operation. Participants believed this facilitated decision-making and hence a smooth evacuation procedure.

##### Feeling united

Participants frequently noted that they had experienced a strong feeling of unity. Some relate bringing out the best in people to crises in general. Others relate this to strong relationships with colleagues but also with authorities.

> *“We, from the Safety Region, are working on getting to know each other outside crisis situations and this led to close contacts, also with the crisis coordinator of the hospital, and it really helped us to do it together*.*” (Participant 7)*

##### Concise communication

There was a need for straightforwardness, as there was no time for extensive discussions and prevarication. Participants appreciated the concise communication, indicating a high level of decisiveness and openness.

> *“*…*there was a lot of transparency; speak to one another if you don’t know or if you feel annoyed, so you can move on*.*” (Participant 5)*

#### 3.2.4 Non-decision-making topics

##### Limitations to NPCC

Although participants positively valued the use of NPCC in the hospital evacuation, there were some limitations mentioned. The majority of the participants referred to the numerous ambulances that had been waiting on the parking lot for several hours before initiating patient transfers to other facilities. Some participants expressed their concern: “…*we have almost 60 ambulances waiting on the parking lot and we are in the midst of the pandemic, so ambulances are needed throughout the country*.., *and secondly, the water was coming, what if we were too late…” (Participant 7)*

Various explanations were provided. Initially, NPCC officials aimed to assign a hospital bed to every patient first, before letting any ambulance depart. This way they tried to ensure that every patient could be transferred to the appropriate facility.

One participant mentioned it took time for NPCC to scale up the capacity that was needed to establish patient acceptance agreements. Another participant believed NPCC requested too many ambulances all at once. Other participants suggested that a lack of knowledge among hospital staff on how to use the NPCC tool to find a hospital bed in another facility contributed to delayed departures of ambulances.

Moreover, predictions on when the water peak would reach the hospital area unexpectedly changed, and the hospital had to be cleared 12 hours earlier than initially planned. According to some, this contributed to the delayed departure of ambulances.

Additionally, it was reported that some ambulances arrived prematurely, due to eagerness to assist in the evacuation operation.

One participant thought the regional safety authority should have been in charge of the evacuation of patients. Especially since NPCC was not designed for, and had no experience in, emergency responses.

> *“If you have more time, don’t get me wrong, then it [NPCC] is the better option because it is much more patient-oriented, more gradual. But if you’re dealing with a crisis then you have to put the regional safety authority in charge*.*” (Participant 11)*

If NPCC were to be used in the future, it was suggested that hospitals arrange trainings together with NPCC and the safety authorities to expand knowledge and expertise.

##### Importance of local and regional business continuity

The hospital evacuation raised awareness amongst participants on consequences for local and regional business continuity in case of emergency events, such as flooding. They addressed the need for planning how to continue regional (emergency) healthcare when a hospital is out of order. *“Do I have a plan for what to do when there is a loss of healthcare? No. Acute loss of care? No. Would it help? I think so. To start thinking about it together*.*” (Participant 7)*

### 3.3 Quantitative results

When the evacuation decision was made, 245 patients were admitted to VCMC. 20 patients were admitted for day-treatments or diagnostics and therefore excluded. One patient died in-hospital before the evacuation commenced and was therefore excluded. Of the remaining 224 patients, 85 (37.9%) were already planned to be discharged on this day. Thirty-one (13.8%) patients were discharged prematurely, and 108 (48.2%) patients were transferred to other hospital facilities. The return transfer of patients to VCMC occurred 5 days after the evacuation event. The flowchart is shown in Figure 4.

**Figure 3.**
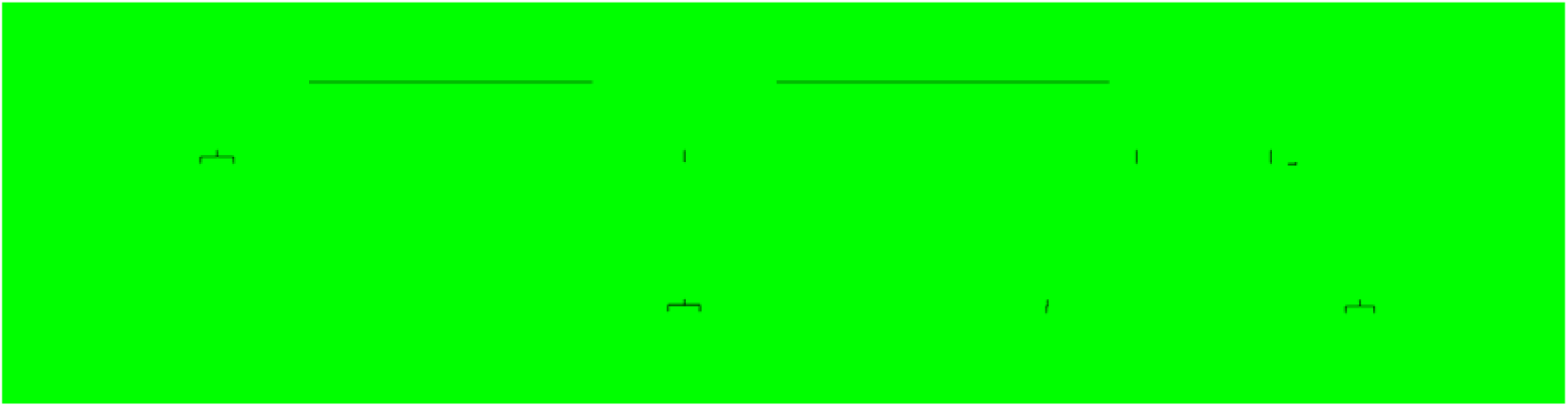
Flowchart of the evacuated patients *CCI: Charlson Comorbidity Index; IQR: Interquartile Range*

No patients died unanticipated during the evacuation. In the 30 days following the evacuation, 8 (3.6%) patients deceased. All had limitations of life-sustaining interventions, indicating a code status do not resuscitate (DNR), do not intubate (DNI) and/or no ICU, and none of the deaths could be directly attributed to the evacuation itself.

Events in prematurely discharged patients and in patients who were transferred to other hospitals are provided in Appendix II.

## 3 DISCUSSION

This mixed-methods study explored the decision-making process of the evacuation of VCMC in Venlo on July 16th 2021 and assessed patients’ mortality and readmission rates to understand the clinical consequences of the evacuation. Three themes were constructed from the interviews; risk-assessment, COVID-19 experience and collaboration. Results showed that the recent experience from the pandemic played a pivotal role in the course of this evacuation event and paved the way for an uneventful evacuation procedure. Furthermore, the use of a national patient coordination center (NPCC) enabled to rapidly assess capacity of potential destination hospitals and to facilitate individual patient transfers. Additional findings showed that the efficiency of large-scale ambulance dispatches could be improved and underlined that business continuity plans should be expanded. No patients died unanticipated during the evacuation and no deaths could be attributed to the evacuation itself.

Only few previous studies reported on evacuation-associated mortality and morbidity [25,26]. Direct comparison is complicated because of differences in the cause of evacuations as well as locoregional circumstances. Risk factors for increased mortality associated with flood-related hospital evacuations previously described include discontinuation of critical services such as oxygen supplies and parenteral feeding, and lack of infrastructures (both internal and external, including communication capabilities, transportation systems and power supply) [25,27]. Another factor associated with mortality is lack of hospital staff to take care of patients [25]. The VCMC evacuation suggest that the evacuation-associated risks to patients were minimal or may be even absent. This could be explained by the fact that during the evacuation internal infrastructures were undamaged and allowed for continuation of critical services as well as internal and external communication. Transportation using NPCC allowed for continuation of healthcare, as ambulances were fully staffed and equipped.

Participants in this study experienced a poor translation of the significance of the threat by the governmental authorities for execution of public works and water management. In a recent evaluation report initiated by the regional safety authority, this issue also came up as point of improvement [28]. According to the report, organizations experienced difficulties to accurately specify water level estimates for some critical locations, including the hospital. This was largely due to the impact of unprecedented rapid rising of the water level in several smaller upstream waterways flowing into the Meuse, which were not incorporated in existing risk-assessment models [28]. Furthermore, the rapid increase of water levels made the impact difficult to predict. The evaluation report concluded that assessments on water levels and weak spots in river banks could be enhanced, thereby improving decision-making [28].

Evacuation decisions are often based on little information [2]. Evaluations of other hospital evacuations show that it is impossible to develop one crisis plan that can be applied to all types of hospital disasters [5]. Some hospitals are more likely to be exposed to certain hazards for geophysical reasons. In the Netherlands for example, floods form a substantial risk [9]. Therefore, infrastructure assessments of hospitals should be performed to estimate the degree of hospital vulnerabilities and the potential consequences of floods for the hospital and adjacent areas, preferably before they are built [29].

The COVID-19 experience played a major role in this particular evacuation, and especially the use of the patient transfer system NPCC was positively valued. It is evident from this study that NPCC enabled to identify real-time capacity in appropriate destination hospitals and hence facilitated patient relocation. To the best of our knowledge, this is the first documented hospital evacuation in which such a national patient coordination center was used. A review on hospital evacuations revealed that the relocation of patients often entails difficulties, especially with regards to inter-facility transport modes [30]. Furthermore, many hospitals are not prepared for large-scale evacuation of vulnerable patients, such as older adults and ICU patients [3]. These difficulties complicate the decision to evacuate a hospital. Some hospitals make patient acceptance agreements in times of no disaster to prepare for future evacuation events. This was the case during the Fukushima nuclear disaster in 2011. Although patient acceptance agreements with neighboring hospitals were in place, these agreements were not found to be effective because the surrounding hospitals were prompted to evacuate as well [25]. The reach of a disaster depends on the disaster type and its magnitude, which – for some natural disasters - complicates future patient disposition planning.

Additional findings from this study showed concerns among participants about ambulances that had been waiting on the parking area for several hours until patients were ready to be transported to other hospitals. A similar situation occurred during the evacuation of another Dutch hospital (VU Medical Center, Amsterdam), when approximately 30 ambulances were waiting while the evacuation procedure had not yet commenced [31]. This phenomenon may be explained by uncertainties regarding the timeframe of evacuation and the willingness amongst ambulance organizations and individual ambulance crews to assist. Although it may provide assurance to hospital organizations, one should realize that ambulances, while waiting for the start of the evacuation, may cause ambulance shortages in a wide region.

Finally, this study again shows a basic principle in hospital disaster preparedness that hospitals should assess their vulnerability and take precautionary measures [32]. These include the safeguarding of critical services and planning for regional loss of healthcare capacity. The findings are in line with previous studies that identified planning for continuity to be crucial in hospital disaster resilience [33].

### 4.1 Strengths and limitations

The research was performed by a researcher who was not related to any of the participants and had no conflicts of interest. The mixed-methods design of this study allowed for gaining insight in the evacuation decision as well as its effect on patient-related outcomes. Although interview guides were agreed with the project team, findings may be influenced by question bias due to the semi-structured approach. The findings may also be influenced by social desirability bias, because of possible interests in the study outcomes among the participants. By anonymizing the collected data, it was attempted to reduce bias. The quantitative data include a relatively large study population, which may reduce bias. This study did not explore patient experiences related to the evacuation. Patient perspectives were, however, beyond the decision-making scope of this study. Due to the increasing worldwide occurrence of natural disasters9, the findings of this study may help to improve the evacuation preparedness of other hospitals.

Overall, it should be noted that this particular evacuation procedure was pre-emptive, unlike some previously described evacuation events [31,33]. The evacuation procedure could be executed with intact hospital services, infrastructure and communication networks.

Furthermore, the flood event did not cause casualties in the Netherlands [34] which limited regional healthcare demands and may have facilitated an uneventful evacuation procedure.

## 4 CONCLUSION

The experiences of the COVID-19 pandemic and the availability of a national patient coordination center were found to be key factors in performing this evacuation. This allowed for the swift identification of available capacity in appropriate destination hospitals. We recommend that hospitals regularly assess potential contingencies and plan for loss of healthcare services.

## Data Availability

All data produced in the present study are available upon reasonable request to the authors

## ABBREVIATIONS

ED: Emergency Department
DNI: Do Not Intubate
DNR: Do Not Resuscitate
IHCDs: Internal Hospital Crises and Disasters
ICU: Intensive Care Unit
IQR: Interquartile Range
NPCC: National Patient Coordination Center
VCMC: VieCuri Medical Center

