## Appendix I for "Addressing a climate emergency amidst the COVID-19 pandemic: A mixed-methods study on a hospital evacuation during the 2021 European floods"

**Appendix I.** Interview guideline- domains and corresponding questions.

Introduction

- Could you describe your role in the evacuation of the hospital?
- Could you describe the decisions you were involved with?

Available information

- Which information did you have at hand?
- To what extent could you use plans and procedures and to what extent did you have to improvise?

Assessing risks

- What was your understanding of the consequences of your decision?
- What risks did you consider at that time?

Previous experience

- To what extent did personal, or previous experiences, play a role in making decisions?

(Un)certainties

- Was it clear to you what everyone’s tasks were?
- Have you ever felt insecure about decisions and why?

Lessons learned

- Looking back, do you wish you had done things differently?
- What do you feel most content with?
