## Appendix II for "Addressing a climate emergency amidst the COVID-19 pandemic: A mixed-methods study on a hospital evacuation during the 2021 European floods"

**Appendix II.** Events in prematurely discharged patients and in patients who were transferred to other hospitals

*Events in prematurely discharged patients*
Of 31 prematurely discharged patients, 3 (9.7%) died within 30 days following discharge. Of these, 2 patients died in a hospice. The transfer from hospital to hospice was planned for these patients, but now occurred some days earlier. The other patient suffered from metastatic cancer and deceased 13 days after evacuation. Three patients (%) were readmitted 5 days after evacuation, when the hospital reopened.

*Events in patients transferred to other hospitals*

Of the 108 patients who had been transferred to other hospitals, 4 patients died (3.7%) within 30 days (day 9, 12, 20 and 26). Of these, 2 patients had untreatable malignancies, 1 had an intracerebral hemorrhage and 1 patient had a severe COVID-19 infection and comorbidity. All of these patients had limitations of life sustaining care (DNR, DNI and NO ICU). One patient was discharged from the receiving facility 2 days post evacuation and readmitted 5 days after evacuation.
